# Interim 2023/24 season influenza vaccine effectiveness in primary and secondary care in the United Kingdom

**DOI:** 10.1101/2024.02.27.24303426

**Authors:** Heather Whitaker, Beth Findlay, Jana Zitha, Rosalind Goudie, Katie Hassell, Josie Evans, Panoraia Kalapotharakou, Utkarsh Agrawal, Beatrix Kele, Mark Hamilton, Catherine Moore, Rachel Byford, Julia Stowe, Chris Robertson, Anastazia Couzens, Gavin Jamie, Katja Hoschler, Kathleen Pheasant, Elizabeth Button, Catherine Quinot, Tim Jones, Sneha Anand, Conall Watson, Nick Andrews, Simon de Lusignan, Maria Zambon, Christopher Williams, Simon Cottrell, Kimberly Marsh, Jim McMenamin, Jamie Lopez Bernal

## Abstract

We report 2023/24 season interim influenza vaccine effectiveness for three studies, including primary care in Great Britain, hospital settings in Scotland and hospital settings in England. Adjusted vaccine effectiveness against all influenzas ranged from 63% (46 to 75%) to 65% (41 to 79%) among children aged 2-17, from 36% (20 to 49%) to 55% (43 to 65%) among adults 18-64 and from 40% (29 to 50%) to 55% (32 to 70%) among adults aged 65 and over.

## Introduction

In the United Kingdom (UK), seasonal influenza vaccination is offered freely to those aged ≥65, aged 16-64 within clinical groups and at increased risk of severe influenza-related outcomes, and children aged 2-15 [1]. In Scotland this offer was extended to healthy adults aged 50-64 [2]. The 2023/24 season northern hemisphere influenza vaccine included an updated A(H1N1)pdm09 strain: a cell culture-propagated A/Wisconsin/67/2022 or egg-propagated A/Victoria/4897/2022 (H1N1)pdm09-like virus, while the A(H3N2) and B components remained the same as for 2022/23 [3].

Throughout the UK, indicators of influenza activity suggested low levels of circulation prior to December 2023 [4-6]. Influenza activity rose throughout December, reaching a modest peak by late December 2023, before dropping slightly early-mid January 2024. However, by late January 2024, influenza activity was increasing again. To January 2024, the UK has primarily seen co-circulation of influenza A(H1N1)pdm09 and A(H3N2).

We report interim vaccine effectiveness in primary and secondary care settings within the UK.

## Methods

We brought together primary care sentinel swabbing data from England, Scotland and Wales (GB-PC) to conduct two separate data-linkage studies on hospitalised patients in England (EN-H) and Scotland (SC-H). Study characteristics are summarised in Table 1. Methods for all three studies have been described elsewhere [7-10]; a protocol for GB-PC is provided in supplementary material.

**Table 1.**
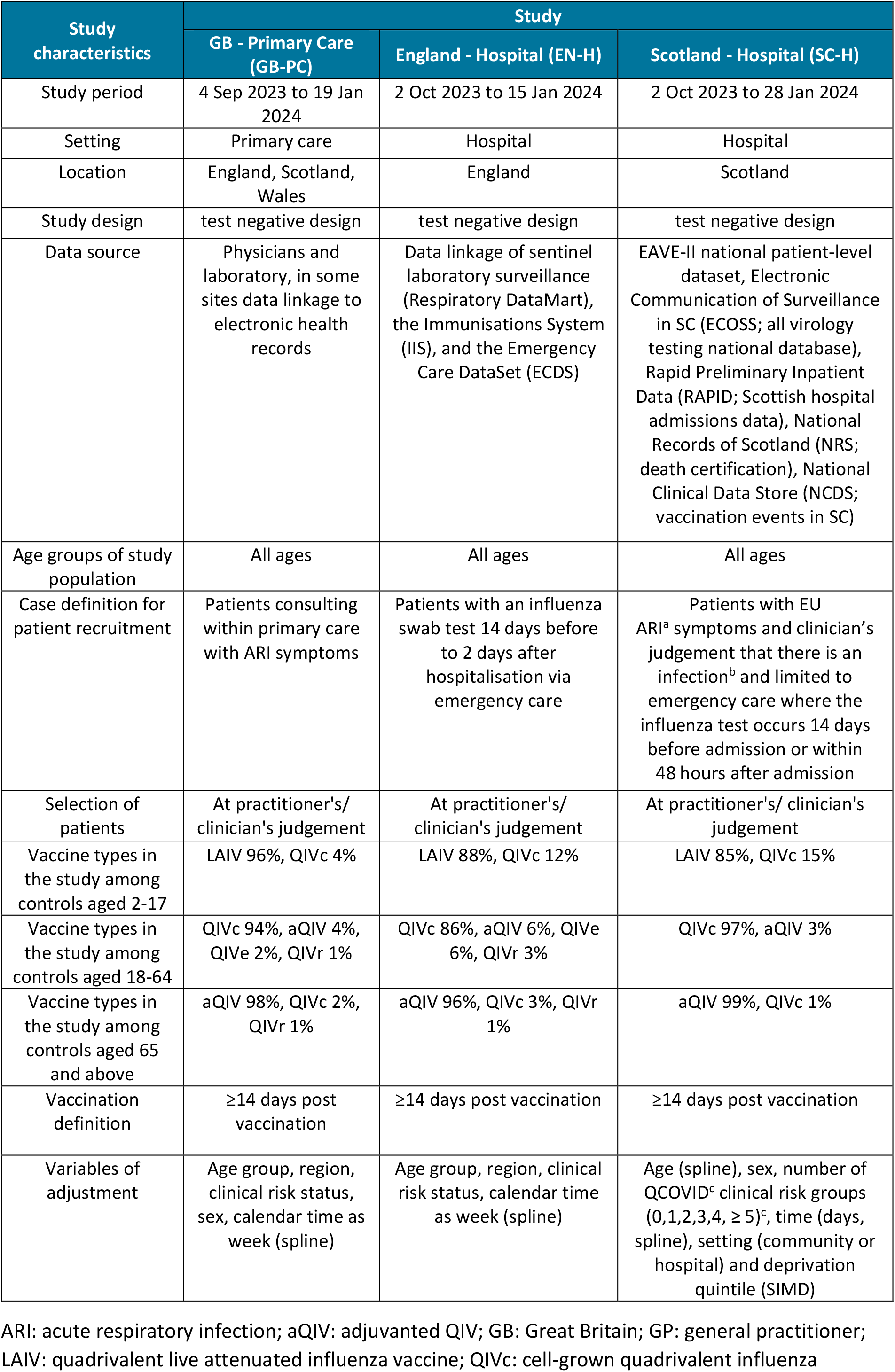
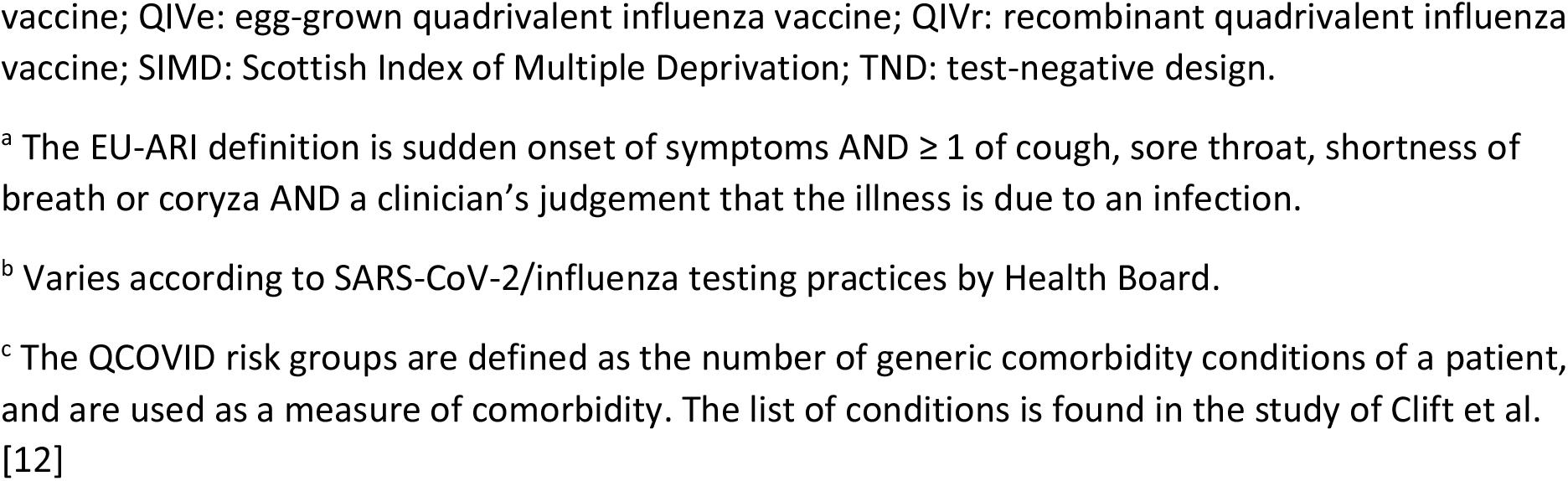
Summary of methods and characteristics of three 2023/24 season influenza vaccine effectiveness studies in Great Britain.

| Study characteristics | Study |  |  |
| --- | --- | --- | --- |
|  | GB - Primary Care (GB-PC) | England - Hospital (EN-H) | Scotland - Hospital (SC-H) |
| Study period | 4 Sep 2023 to 19 Jan 2024 | 2 Oct 2023 to 15 Jan 2024 | 2 Oct 2023 to 28 Jan 2024 |
| Setting | Primary care | Hospital | Hospital |
| Location | England, Scotland, Wales | England | Scotland |
| Study design | test negative design | test negative design | test negative design |
| Data source | Physicians and laboratory, in some sites data linkage to electronic health records | Data linkage of sentinel laboratory surveillance (Respiratory DataMart), the Immunisations System (IIS), and the Emergency Care DataSet (ECDS) | EAVE-II national patient-level dataset, Electronic Communication of Surveillance in SC (ECOSS; all virology testing national database), Rapid Preliminary Inpatient Data (RAPID; Scottish hospital admissions data), National Records of Scotland (NRS; death certification), National Clinical Data Store (NCDS; vaccination events in SC) |
| Age groups of study population | All ages | All ages | All ages |
| Case definition for patient recruitment | Patients consulting within primary care with ARI symptoms | Patients with an influenza swab test 14 days before to 2 days after hospitalisation via emergency care | Patients with EU ARI<sup>a</sup> symptoms and clinician's judgement that there is an infection<sup>b</sup> and limited to emergency care where the influenza test occurs 14 days before admission or within 48 hours after admission |
| Selection of patients | At practitioner's/ clinician's judgement | At practitioner's/ clinician's judgement | At practitioner's/ clinician's judgement |
| Vaccine types in the study among controls aged 2-17 | LAIV 96%, QIVc 4% | LAIV 88%, QIVc 12% | LAIV 85%, QIVc 15% |
| Vaccine types in the study among controls aged 18-64 | QIVc 94%, aQIV 4%, QIVe 2%, QIVr 1% | QIVc 86%, aQIV 6%, QIVe 6%, QIVr 3% | QIVc 97%, aQIV 3% |
| Vaccine types in the study among controls aged 65 and above | aQIV 98%, QIVc 2%, QIVr 1% | aQIV 96%, QIVc 3%, QIVr 1% | aQIV 99%, QIVc 1% |
| Vaccination definition | ≥14 days post vaccination | ≥14 days post vaccination | ≥14 days post vaccination |
| Variables of adjustment | Age group, region, clinical risk status, sex, calendar time as week (spline) | Age group, region, clinical risk status, calendar time as week (spline) | Age (spline), sex, number of QCOVID<sup>c</sup> clinical risk groups (0,1,2,3,4, ≥ 5)<sup>c</sup>, time (days, spline), setting (community or hospital) and deprivation quintile (SIMD) |

ARI: acute respiratory infection; aQIV: adjuvanted QIV; GB: Great Britain; GP: general practitioner; LAIV: quadrivalent live attenuated influenza vaccine; QIVc: cell-grown quadrivalent influenza
vaccine; QIVe: egg-grown quadrivalent influenza vaccine; QIVr: recombinant quadrivalent influenza vaccine; SIMD: Scottish Index of Multiple Deprivation; TND: test-negative design.
<sup>a</sup> The EU-ARI definition is sudden onset of symptoms AND $\geq 1$ of cough, sore throat, shortness of breath or coryza AND a clinician's judgement that the illness is due to an infection.
<sup>b</sup> Varies according to SARS-CoV-2/influenza testing practices by Health Board.
<sup>c</sup> The QCOVID risk groups are defined as the number of generic comorbidity conditions of a patient, and are used as a measure of comorbidity. The list of conditions is found in the study of Clift et al. [12]

All studies used the test negative design, applying logistic regression with adjustments as outlined in Table 1 to estimate the odds ratio (OR) of vaccination in influenza positive cases and influenza negative controls. Adjusted vaccine effectiveness (aVE) is reported as a percentage: 100×(1-OR). For the GB-PC study, patients presented to primary care with acute respiratory infection (ARI). In EN-H, influenza A subtype VE analyses were restricted to data from labs that carried out subtyping. In SC-H, potential vaccine contaminants and influenza co-detections were removed from all analyses. SARS-CoV-2 positive controls were excluded in the GB-PC and EN-H studies [11].

A sample of influenza virus positive primary care specimens were further characterised, as described in supplementary material.

## Results

The GB-PC study included 1,193 cases and 12,098 controls, including 6,343 samples from England, 6,108 from Scotland and 840 from Wales; the number of specimens positive for influenza A(H1N1)pdm09 was 461, 475 A(H3N2), 215 influenza A (untyped) and 46 influenza B, with 4 dual infections. The EN-H study included 1,359 cases and 22,539 controls; 770 influenza A samples were untyped, 161 were influenza A(H1N1)pdm09, 395 were influenza A(H3N2) (note that more laboratories subtyped influenza A/H3 than A/H1), 32 were influenza B, including 5 dual infections. The SC-H study included 1,977 cases and 34,476 controls; 1,567 influenza A samples were untyped, 172 were influenza A(H1N1)pdm09, 188 were influenza A(H3N2), and 50 were influenza B.

We note from Table 1 the distribution of influenza vaccine types among controls. Those aged 2-17 primarily received live attenuated influenza vaccine (LAIV) via nasal spray, those aged 18-64 mostly received quadrivalent cell-based vaccine (QIVc) and those aged ≥65 mostly received adjuvanted egg-based vaccine (aQIV). Use of other vaccine types—standard-dose quadrivalent egg-based vaccines (QIVe) and recombinant quadrivalent vaccines (QIVr)—was limited.

Vaccine effectiveness for all studies is summarised in Figure 1. There were insufficient influenza B samples in all studies to estimate VE against influenza B.

**Figure 1.**
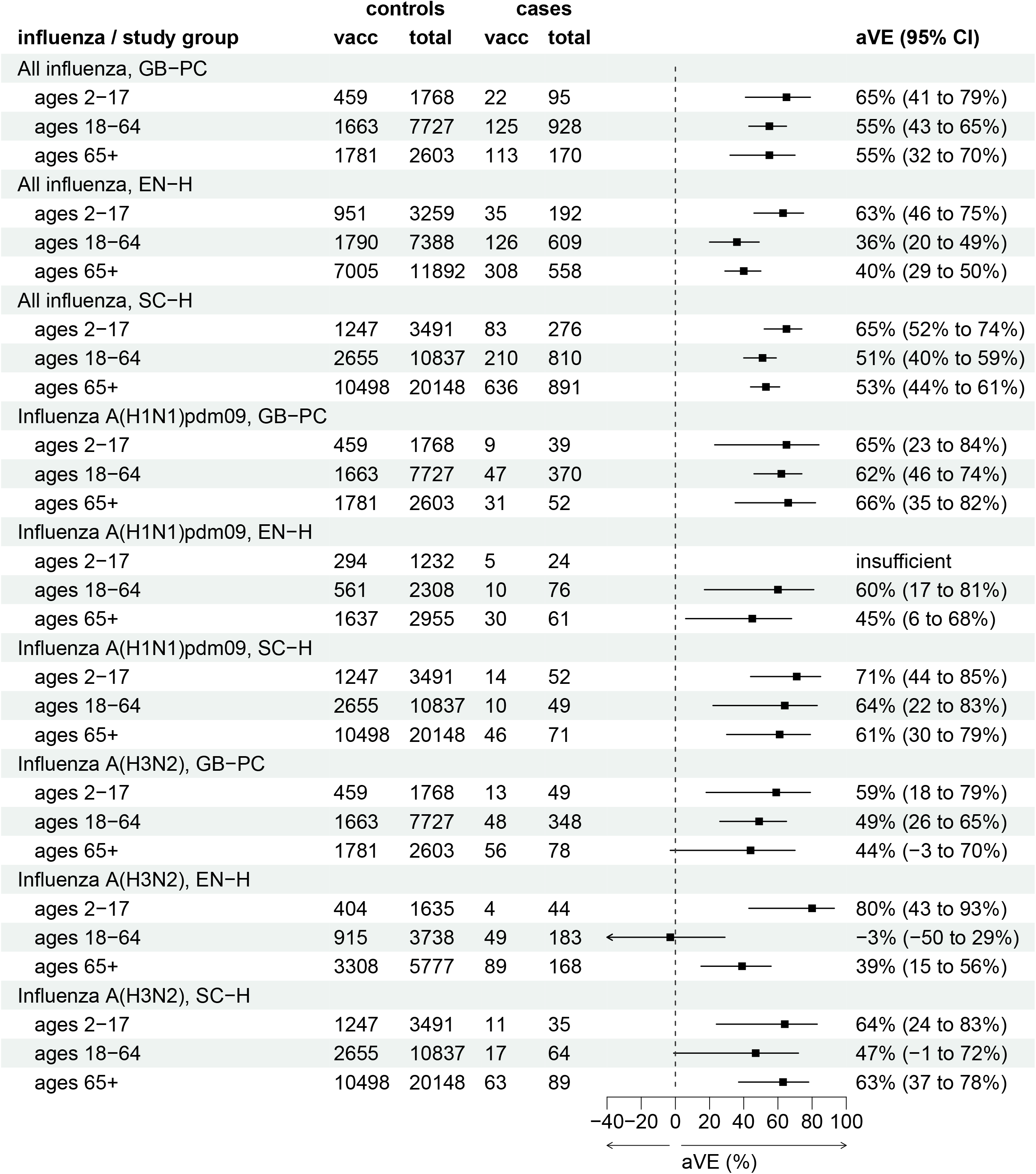
Interim adjusted vaccine effectiveness (aVE) against all laboratory-confirmed influenza (A and B), A(H1N1)pdm09 and A(H3N2), by each of three studies in Great Britain and by age group, influenza season 2023/24

For all influenza cases (both influenza A and B) across all settings, aVE ranged from 63% (95% Confidence Interval (CI) 46 to 75%) to 65% (95% CI: 41 to 79%) among children aged 2-17, from 36% (95% CI: 20 to 49%) to 55% (95% CI 43 to 65%) among adults 18-64, and from 40% (95% CI: 29 to 50%) to 55% (95% CI: 32 to 70%) among adults aged 65 and over.

In all settings, aVE against influenza A(H1N1)pdm09 ranged from 65% (95% CI: 23 to 84%) to 71% (95% CI: 44 to 85%) among children aged 2-17, from 60% (95% CI: 17 to 81%) to 64% (95% CI: 22 to 83%) among adults aged 18-64 and from 45% (95% CI: 6 to 68%) to 66% (95% CI: 35 to 82%) among adults aged 65 and above.

aVE against influenza A(H3N2) ranged from 59% (95% CI: 18 to 79%) to 80% (95% CI: 43 to 93%) among children aged 2-17, from -3% (95% CI: -50 to 29%) to 49% (95% CI: 26 to 65%) among adults aged 18-64 and from 39% (95% CI: 15 to 56%) to 63% (95% CI: 37 to 78%) among adults aged 65 and over. Except for one result (SC-H, ages 65+), aVE point estimates against influenza A(H3N2) aVE were lower than those against influenza A(H1N1)pdm09.

Where influenza viruses from positive specimens in the GB-PC study were further characterised, all A(H3N2) viruses belonged in genetic subclade 3C.2a1b.2a.2 in the 2a.3a.1 subgroup, while A(H1N1)pdm09 viruses were split between subgroups 6B.1A.5a.2a (70%) and 6B.1A.5a.2a.1 (30%).

## Discussion

During a period of co-circulation of influenza A(H1N1)pdm09 and A(H3N2) in the UK we found evidence of moderate vaccine effectiveness in both children and adults. Our results concur with those reported for Canada during the early 2023/24 season, who reported moderate vaccine effectiveness during an influenza A(H1N1)pdm09-dominated period [12].

During the 2022/23 season, estimates of interim vaccine effectiveness against influenza A(H1N1)pdm09 in Europe, including some corresponding results for EN-H and SC-H, were typically lower than those we report for the 2023/24 season, especially among those aged 65 and above [9]. Our results suggest that vaccine effectiveness may have been improved by the update to the A(H1N1)pdm09 component of the northern hemisphere vaccine for 2023/24.

The 2023/24 A(H3N2) northern hemisphere vaccine strain belongs to subgroup 2a of genetic subclade 3C.2a1b.2a.2 Experiments to evaluate the vaccine’s ability to recognise subgroup 2a.3a.1 viruses similar to those circulating have reported mixed findings [14]. Our 2023/24 vaccine effectiveness estimates against A(H3N2) are higher than those reported for 2022/23 in SC-H as well as among children and adults aged 65 and above in EN-H [7], suggesting that the differences in subgroup of the vaccine and circulating viruses have not resulted in reduced VE. However, no effectiveness was demonstrated in the 2023/24 EN-H for those aged 18-64 years. Healthy adults aged 50-64 were influenza vaccine eligible in 2022/23, but in 2023/24 this was withdrawn in England and Wales while continuing in Scotland; evidence for residual protection leading to reduced VE will be investigated at the end of the season.

Interim estimates are inevitably restricted by sample size. End-of-season final estimates will further explore VE by vaccine type and possibly provide VE against influenza B, depending on circulation during the rest of the season. VE estimates were often lower in EN-H; hospitalisation definitions differ to SC-H and further exploration of this is planned. In a hospital setting, testing is among patients presenting with ARI symptoms, but a limitation is that this may not be the main reason for hospitalisation. Unlike previous seasons, Scotland successfully collected individual-level vaccination data for children from all health boards, resulting for the first time in a national hospital study population.

Estimates of interim 2023/24 season influenza vaccine effectiveness within the UK are encouraging, especially in that reasonably robust effectiveness against hospitalisation with both influenza A(H1N1)pdm09 and A(H3N2) was demonstrated among those aged 65. These findings further reinforce the importance of the annual UK influenza vaccination offer.

## Supporting information

Supplementary material GB-PC protocol

## Data Availability

Due to legal reasons supporting data is not available to share.

## Ethical statement

UK public health agencies have permission to process patient confidential information for national surveillance of communicable diseases under: Regulation 3 of the Health Service Regulation 2002 for England, the Public Health (Scotland) Act 2008 and the NHS Scotland Act 1978 for Scotland, and the Public Health Wales National Health Service Trust (Establishment) Order 2009 for Wales. As such, specific ethical approval was not necessary.

