## Supplementary material GB-PC protocol for "Interim 2023/24 season influenza vaccine effectiveness in primary and secondary care in the United Kingdom"

### Surveillance of respiratory infections and vaccine effectiveness using primary care sentinel networks in the United Kingdom

### 2023/24 Proposal

Version 1.0 04/07/2023

Version 1.1 25/09/2023

Version 1.2 16/02/2024 updated laboratory methods

### 1) Background

#### General

Because influenza viruses constantly evolve and vaccines are reformulated every year, vaccine effectiveness (VE) estimates from previous years cannot be used to estimate VE in the subsequent years. Having annual influenza VE estimates as soon as possible after the start of a seasonal influenza epidemic or pandemic and monitoring it along its course is essential to:

- decide on recommendations for the use of the vaccine
- target complementary or alternative public health measures (e.g. antivirals) for population segments where vaccine is less effective.
- trigger further investigations on seasonal and pandemic vaccines (improve composition, use of adjuvants, need for booster doses).

The UK introduced a universal childhood influenza vaccine programme targeted for 2-16 year old children from 2013-14. In summer 2016, the US Advisory Committee on Immunisation Practice recommended the temporary suspension of use of LAIV in the USA following the CDC assessment of the lack of effectiveness of LAIV in children in the 2015-16 season, with little evidence of significant VE; the suspension was rescinded in spring 2018. Although these results are at odds with those seen in several other countries using LAIV, including the UK, it will be important to ensure the evaluation of the effectiveness of LAIV in children.

In recent seasons, it has become apparent that the current inactivated influenza vaccine programme provides sub-optimal protection to the age-groups >65 years of age, especially against influenza A/H3N2. Adjuvanted vaccines have been preferentially recommended since the 2018/19 season to all in this age-group. Since 2019/20, cell-based quadrivalent vaccines have also been available and used preferentially over standard-dose egg-based vaccines. The recombinant vaccine has also been available since 2021/22, though uptake has been low. In addition, all adults 50-64 years of age have been eligible to receive either cell or egg-based quadrivalent influenza vaccines since the 2020/21 season with the aim of reducing pressure on the health service during the COVID-19 pandemic. It is thus critically important to obtain accurate estimates of the relative effectiveness of these influenza vaccines in the relevant age groups targeted with different vaccine types (2-17, 18-64, 65+), and bringing together data from across the UK will help this aim. This proposal concerns the use of the test-negative case control method to estimate influenza vaccine effectiveness using the various UK sentinel GP swabbing schemes, with a particular focus on the absolute and relative effectiveness of these vaccines.

Further, some influenza positive sentinel swabs are targeted for genetic characterisation. Exploring vaccine effectiveness by sub-clade may help us better understand the impact of antigenic drift on vaccine effectiveness and inform vaccine strain selection. Bringing together this information from sentinel schemes across the UK helps maximise our potential to produce vaccine effectiveness estimates by sub-clade and optimize use of this valuable data.

Finally a number of RSV treatment strategies are approaching. There is a need to establish systems to monitor the baseline epidemiology of RSV prior to introduction of these and for this to then provide a platform to monitor the RSV vaccine programme impact and effectiveness. The burden of disease due to RSV is increasingly being recognised – particularly in young infants and also potentially the elderly, though questions remain. There is a need to establish RSV disease burden in the UK, particularly in younger children and the elderly to inform optimal future use of these new vaccines and to provide a baseline for subsequent impact studies stage.

### *2) Aim and Objectives*

#### 2.1 Aim

For the 2023/24 season, the aims are to:

- Estimate the pooled influenza vaccine effectiveness for laboratory confirmed infection in primary care across three age groups: children aged 2-17, working-age adults 18-64 and the elderly aged 65+ in the UK.
- Monitor RSV epidemiology in primary care with a focus on children <5 years of age

#### **2.2** Primary objectives

The primary objective is to estimate 2023/24 seasonal influenza vaccine effectiveness (VE):

- Overall for any flu type,
- by influenza type (A or B),
- by influenza subtype (A/H1 or A/H3);

within each of 3 broad age groups:

- 2-17 years (and stratified by nasal vs quadrivalent inactivated vaccine),
- 18-64 years of age (and stratified by QIVc vs QIVr vs QIVe),

- >65 years of age (and stratified by aQIV and QIVc).

To describe the epidemiology of RSV in children.

#### 2.3 Secondary objectives

To estimate:

- VE by risk status
- VE by influenza main clades/sub-clades
- VE by time since vaccination
- VE by period (depending on circulation patterns)
- VE by prior vaccination status in 2022/23 overall and by type/sub-type (England, Scotland)
- The interaction of age and VE in adults age 18+
- VE against asthma/COPD exacerbation (England only)

### *3) Methods*

#### 3.1 Study designs

- Test-negative case-control (TNCC) design will be used to estimate VE.
- For RSV descriptive statistics on swab positivity will be collated, by age group, gender, month and risk status.

#### 3.2 Study population and data source

***Definitions***

The study will be undertaken in the registered population of sentinel general practice surveillance networks across the UK that undertake respiratory swabbing, details of which are outlined. The surveillance schemes are: Royal College of General Practitioners (RCGP) Research and Surveillance Centre (RSC), Public Health Wales Communicable Disease Surveillance Centre Sentinel GP Surveillance, Public Health Scotland (PHS) CARI surveillance programme.

The study population will be patients presenting to their general practice (GP) or other primary and community care based acute respiratory infection (ARI) facility during the 2023/24 influenza season study period with an acute respiratory illness fitting the case definition (influenza-like illness [ILI] or an acute respiratory infection [ARI]). A registered patient will be fully registered (not temporary) with a valid NHS or CHI number and no qualification period.

Cases will be patients who test positive for influenza A or B virus by real-time PCR.

Controls will be patients with the same symptoms who tested negative for influenza A and B, and negative for SARS-CoV-2.

***Study Site Settings***

| |
|---|
| *English RCGP RSC and UKHSA sentinel system:*<br>*Patients who contacted their GP, where the GP (or other community facility) that was part of the Royal College of General Practitioners (RCGP) Research and Surveillance Centre (RSC) sentinel network; and who had a sentinel respiratory swab taken or conducted a self-swab with a result for influenza during the 2023/24 influenza season. Indications for swabbing are influenza-like illness, COVID19-like illness, or another acute respiratory infection including exacerbations of asthma/COPD with onset within the last 10 days. Patients are either consented and sampled in their practice as part of a face-to-face consultation, or register online for a self-swab using a voucher code through takeatestuk.com. All sentinel swabs are tested for respiratory infections including influenza and RSV using RT-PCR at UKHSA's Virus Reference Laboratory. Vaccination data are available via linkage to GP records and NIMS.*<br>*The RSC is in the process of moving sentinel sample requests and delivery into the computerised pathology laboratory links system in the coming season. This will remove the need for data transcription in the laboratory and automatically transfer results back into general practice computerised medical records and to patients via the NHS App.* |

| |
|---|
| *Scottish PHS scheme:*<br>*Patients who contacted their GP, where the GP was one of around 140 sentinel practices taking part in the CARI surveillance programme, and who had a respiratory swab taken with a result for influenza during the 2022/2023 influenza season. Indications for swabbing were acute respiratory infection (ARI) with sudden onset within the last 7 days, and aligning with the ECDC definition of ARI. Patients are either consented and swabbed by their GP face-to-face, or they use a self-swabbing kit to take the swab themselves. All sentinel swabs are multiplex tested for 10 respiratory infections at the West of Scotland Specialist Virology Centre and results are held by ECOSS (national* |

*database of all virology test results). Vaccination data are available for adults only from the Vaccine Management Tool (VMT). All date can be record-linked using the CHI number.*

*Welsh scheme:*

*Patients who contacted their GP, where the GP was part of the Public Health Wales Sentinel GP Sentinel GP Surveillance Network, and who had a sentinel respiratory swab taken with a result for influenza during the 2022/23 influenza season. Indications for swabbing are influenza-like illness, COVID19-like illness or an acute respiratory infection with onset within the last 10 days. Patients consulting in person are swabbed and consented by their GP as part of a face-to-face consultation or provided with a postal self-swab kit by GPs if consulting via telephone. All sentinel swabs are tested for respiratory infections including influenza and RSV using RT-PCR at Public Health Wales Specialist Virology Centre. Vaccination status is provided by the GP at time of consultation, either sourced from the patients' medical records or self-reported by the patient to the GP.*

#### 3.3 Study period

The start of the influenza vaccination campaign is usually in mid-September as soon as influenza vaccine becomes available. It is theoretically available until supplies run out, although the bulk is delivered in late September through to November. The bulk of childhood vaccinations in Primary schools are delivered October to December.

The study period will commence from 4 September 2023 (the point when influenza vaccination rollout and influenza and RSV transmission may begin) and continue for the duration of the UK influenza season. An early calculation of VE will be performed prior to the February 2024 WHO strain selection committee meeting, with a full analysis carried out at the end of the season.

This proposal relates to an ongoing programme of disease surveillance. The end of the influenza season is not fixed *a priori*. The period of influenza activity is determined using data from the national influenza surveillance system.

RSV statistics will be collated at the end of the influenza season using the same season definition as for influenza.

#### 3.4 VE exposure

***Definition***

Patients will be defined as vaccinated if they have received the 2022/23 seasonal vaccine at least 14 days before the date of the swab. Note that 2 doses of LAIV are recommended for some children, but will be considered vaccinated from 14 days after the first dose.

***Ascertainment***

Vaccination history is based on electronic health records, which may be supplemented by vaccination information given on the swabbing form if available. Vaccination dates are required, ideally accompanied by vaccine type (LAIV-intranasal; injectable-QIVe, injectable-cell-based QIVc, injectable-egg-based QIVe injectable recombinant QIVr and adjuvanted injectable-aQIV). In primary care where possible we encourage recording of brand of vaccine and batch number, which enables vaccine type to be derived.

#### 3.5 VE outcome(s)

The PHS definition of ARI (sudden onset within last 7 days) is –

- Sudden onset of symptoms
- At least one of the following four respiratory symptoms:
    - Cough
    - Sore throat
    - Shortness of breath
    - Coryza

AND

- A clinician's judgement that the illness is due to an infection.

RCGP RSC's ARI phenotype has four sub-groups, and will include events with a sudden onset within 10 days:

- influenza-like illness (ILI),
- exacerbations of chronic lung disease (ECLD),
- lower respiratory tract infection (LRTI), and
- upper respiratory tract infection (URTI).

definition of influenza-like illness:

- An acute respiratory infection (ARI)
- With measured or clinically plausible temperature ≥ 38°C (other than in older people who can have infections without a fever)
- Cough
- Sudden onset and in the absence of a more plausible diagnosis.

The RCGP and Welsh scheme suggest swabbing with onset within the last 10 days, this was extended during the COVID-19 pandemic. However, influenza positivity is highest within the first 7 days, so analyses will aim to restrict to the 0-7 day post onset window.

For the 2023/24 season RCGP are broadening their definition for swabbing to include exacerbations of chronic lung disease such as asthma and COPD. These swabs will be excluded from main analyses (where known), and explored in secondary analyses *if* there are sufficient swab numbers.

Cases are defined as patients with a positive test for influenza, while those with a negative test for influenza and SARS-CoV-2 (but who could have tested positive for other pathogens) are classified as controls.

#### 3.6 VE exclusion criteria

Registered patients may be excluded if they have expressed a wish to be excluded from the surveillance programme; or opted out of sharing their data. However, those who consent to a virology swab will include in their consent record sharing for this purpose.

We ideally look for swabs to fall within 0-7 days of reported illness onset. However, patients will not be excluded where no onset date is given or if the number of days between onset and swab exceeds 7 days (the impact of missing and longer onsets will be explored and multiple imputation methods used). Swabs with implausible onset dates post sample receipt will be retained and the onset date replaced as missing.

Patients must have both a negative influenza A and an influenza B result present to be included as a control.

Patients with nonsensical vaccination histories (e.g. outside the flu season, adults that received LAIV, children that received aQIV) or swab dates (e.g. if swabbed after date of receipt) will be excluded, as will swabs received >21days after the reported date taken.

The study will include only those with known age between 2 and 105 and known sex.

Patients residing outside the boundaries of their respective schemes will be excluded i.e. RCGP/UKHSA patients must be resident in England, PHS patients must be resident in Scotland.

Patients with unknown vaccination status will be excluded. This will include patients with no linkage to either NIMS or a GP record in England, and patients for whom no definite yes/no response to the question on vaccination status was recorded on the Welsh swabbing form. Scottish patients will be assumed to be unvaccinated if no record of vaccination is found.

LAIV-eligible patients aged 2-17 will be excluded if they were vaccinated 0-13 days before symptom onset, or if no vaccination date is available, to ensure positives are not related to LAIV. Patients aged 18+ will not be considered fully vaccinated until 14-days post vaccination. Those swabbed during days 1-13 post vaccination will be included in analyses and assigned a separate exposure category for recent vaccination. Vaccinated adults aged 18+ will not be excluded if no vaccination date is available, instead a multiple imputation approach will be used.

***De-duplication***

It is important to ensure that each individual positive episode is included only once, but given that a patient may experience multiple influenza episodes each season, patients may contribute more than one swab to the study, if swabs can reasonably be considered independent.

Patients who had had more than one test can be included as cases and controls more than once, unless the tests were within 28 days. For repeat tests within 28 days, the second test will be excluded, unless a positive test came after a negative test, in which case the negative result will be excluded.

#### 3.7 Sub-groups

***Risk groups***

Study subjects are categorised according to Department of Health Green Book defined risk categories for influenza vaccination. High risk is determined by the presence of well recognised risk morbidities recorded in the electronic health record for the patient concerned.

- Underlying heart disease
- Chronic respiratory disease
- Diabetes mellitus
- Chronic kidney disease
- Chronic neurological disease
- Splenic dysfunction
- Immunosuppression
- Morbid obesity (BMI>40)
- Chronic liver disease

***Currently pregnant***

- Pregnancy

***Previous vaccination***

Influenza vaccination in the previous season (2022/23)

***Virus genetic characterisation***

Virus clade, and for influenza B lineage

#### 3.8 Data items

Most data items are available through electronic health or laboratory records. Some data items are collected by the consulting clinician in their GP practice (or by individuals if self-swabbing) and entered onto a swab questionnaire.

**Table 1: Data items requested**

| Column | Responses | Explanation |
|---|---|---|
| **Year** | | Year of sample |
| **Lab_Number** | | Local unique laboratory number |

| | | |
|---|---|---|
| **RECEIPT_DATE** | dd/mm/yyyy | Date of sample receipt by lab |
| **SAMPLE_DATE** | dd/mm/yyyy | Date sample taken |
| **ONSET_DATE** | dd/mm/yyyy | Date of onset of illness |
| **PRESENCE_OF_FEVER** | #inp | Not completed on form |
| | No | No |
| | Not known | Not known |
| | Yes | Yes - fever >38 degrees with onset in previous 7 days |
| **PRESENCE_OF_COUGH** | #inp | Not completed on form |
| | No | No |
| | Not known | Not known |
| | Yes | Yes - cough with onset in previous 7 days |
| **PRESENCE_OF_LOSS_OF_TASTE_OR_SMELL** | #inp | Not completed on form |
| | No | No |
| | Not known | Not known |
| | Yes | Yes - cough with onset in previous 7 days |
| **PRESENCE_OF_SHORTNESS OF BREATH** | #inp | Not completed on form |
| | No | No |
| | Not known | Not known |
| | Yes | Yes - SOB with onset in previous 7 days |
| **PRESENCE_OF_WHEEZE (≤5 year)** | #inp | Not completed on form |
| | No | No |
| | Not known | Not known |
| | Yes | Yes - wheeze with onset in previous 7 days |
| **Individual_ID** | | Pseudonomised Individual Identifier |
| **SEX** | m | male |
| | f | female |
| | nk | non known |
| **DATE_OF_BIRTH** | dd/mm/yyyy | Date of birth |
| **CMO_RISKGRP** | #inp | Not completed on form |
| | No | No |
| | Not known | Not known |
| | Yes | Yes |
| **Seasonal_Vaccine_date_23_24** | dd/mm/yyyy | Date of vaccination |
| **Seasonal_Vaccination_23_24** | #inp | Not completed on form |
| | Yes | Vaccinated |

| | | |
|---|---|---|
| | nk | Not known |
| | No | Not vaccinated |

| | | |
|---|---|---|
| **Vaccination_Route_23_24** | #inp | Not completed on form |
| | Intramusucl ar | Vaccinated |
| | Intranasal | Not known |
| | nk | Not known |

| | | |
|---|---|---|
| **Vaccine_Type_23_24** | LAIV | LAIV |
| | aQIV | QIV - adjuvanted |
| | QIVr | QIV - recombinant |
| | QIVe | QIV - egg based |
| | QIVc | QIV - cell based |
| | nk | Not known |
| | #inp | Not completed on form |

| | | |
|---|---|---|
| **Past_Seasonal_Vaccination_22_23** | #inp | Not completed on form |
| | nk | Not known |
| | Yes | Vaccinated |
| | No | Not vaccinated |

| | | |
|---|---|---|
| **COVID19_Autumn23_Vaccine_Date** | dd/mm/yyyy | Date of any COVID19 vaccination since Sept 2023 |

| | | |
|---|---|---|
| **COVID19_Autumn23_Vaccination** | #inp | Not completed on form |
| | nk | Not known |
| | Yes | Vaccinated since Sept 2023 |
| | No | Not vaccinated |

| | | |
|---|---|---|
| **FLU_PCR_Result** | #flub | Influenza B |
| | #h1 | Influenza AH1 |
| | #h3 | Influenza AH3 |
| | #flua | Influenza A(unknown) |
| | #h1h3 | Influenza AH1 AND influenza AH3 |
| | #h1h3flub | Influenza AH1 AND influenza AH3 AND influenza B |
| | #h3flub | Influenza AH3 AND influenza B |
| | #h1flub | Influenza AH1 AND influenza B |
| | #laiv | LAIV strains detected |
| | #nvd | No virus detected |

| | | |
|---|---|---|
| **Other_PCR_Result** | #covid | SARS-CoV-2 |
| | #hmpv | Human Metapneumavirus |
| | #ra | RSV A |
| | #rb | RSV B |
| | #rarb | Both RSV A and RSV B detected |
| | #rsv | RSV untyped |
| | #adeno | Adenovirus |
| | #rhino | Rhinovirus |
| | #corona | Seasonal coronavirus |

| | #nvd | No virus detected |
|---|---|---|

| | | |
|---|---|---|
| **Final_Overall_Result** | #b,#nvd | Influenza B |
| | #h3,#nvd | Influenza AH3 |
| | #h1,#nvd | Influenza AH1 |
| | #nvd,#covid | SARS-CoV-2 |
| | #nvd,#hmpv | Human Metapneumavirus |
| | #nvd,#ra | RSV A |
| | #nvd,#rb | RSV B |
| | #nvd,#rarb | Both RSV A and RSV B detected |
| | #nvd | No virus detected |
| | Results awaited | Results awaited |

Some of the above data items above are not essential to the main study, such as symptom information, these may be used if there is specific interest in symptom groups or to exclude those with COVID19-like illness. The level of detail could be lessened to enable data sharing, e.g. age on 1st September 2023 can be given instead of DOB, indication that vaccination was given ≥0, 14 and 21 days prior rather than precise vaccination dates, lab number can be pseudonymised.

Further genetic characterisation data is requested, where available, for the 2023/24 season circulating strains of interest include:

| genetic group | Like virus | amino acid substitutions |
|---|---|---|
| H1N1 | | |
| 6B.1A.5a.2a | A/Sydney/5/2021 | K54Q, A186T, Q189E, E224A, R259K, K308R |
| 6B.1A.5a.2a.1 | A/Victoria/4897/2022 (vaccine) | P137S, K142R, D260E |
| H3N2 | | |
| 3C.2a1b.2a.2a | A/Darwin/9/2021 (vaccine) | H156S |
| 3C.2a1b.2a.2a.1 | A/Slovenia/8720/2022 | D53G, D104G, K276R |
| 3C.2a1b.2a.2a.1b | A/Catalonia/NSVH161512067/2022 | I140K, R299K |
| 3C.2a1b.2a.2a.3 | A/Norway/24873/2021 | D53N, N96S, I192F |
| 3C.2a1b.2a.2a.3a | | E50K |
| 3C.2a1b.2a.2a.3a.1 | | I140K |
| 3C.2a1b.2a.2a.3b | | I140M |
| 3C.2a1b.2a.2b | A/Thuringen/10/2022 | E50K, F79V, I140K |
| B | | |
| V1A.3 | | |
| V1A.3a.1 | | |
| V1A.3a.2 | B/Austria/1359417/2021 (vaccine) | A127T, P144L, K302R |

#### 3.9 Laboratory methods

***Specimen collection***

GPs take combined throat and nose swabs which will be sent from sentinel GP surveillance networks to the usual laboratory.

***Tests used***

Influenza laboratory confirmation will be undertaken using comparable real-time PCR assays. Samples undergo a molecular analysis for circulating influenza A and influenza B viruses, respiratory syncytial viruses A and B, human metapneumoviruses A and B and COVID-19Further influenza genetic characterisation work is undertaken to determine genetic clade, and for influenza B viruses the lineage.

Suitable influenza positive samples were further characterised by next-generation sequencing of the haemagglutinin (HA) genes of influenza A(H1N1)pdm09, A(H3N2) and influenza B, based on PCR detection cycle threshold (Ct) values ≤ 32, respectively. All genetic characterisation data was generated by the UKHSA Respiratory Virus Reference Unit using full genome amplification protocols for influenza A and B for sequencing on an Illumina MiSeq or Illumina NextSeq instruments. Influenza virus genomes were assembled using an in-house developed pipeline, genetic clade assignments were done using an in-house developed script and confirmed using FluSurver. All influenza whole genomes and HA-only or HA-NA partial genomes were uploaded to GisAID(18).

#### 3.10 Statistical Analysis

Stata 14/17 (StataCorp, College Station, TX, USA) will be used for this data analysis.

***Flu and RSV descriptive analyses***

Cases and non-cases will be described according to:

- Month of swab – count (%)
- Sex – count (%)
- Age group – count (%)
- Region - count (%)
- risk status – count (%)
- Vaccination Status including Vaccination Type – count (%)
- Past season vaccination status – count (%)
- COVID-19 vaccination status – count (%)

Differences in swab positivity within these groups will be tested using Chi-squared or Fisher's exact test, as appropriate. Swabs with a gap from onset to swab >7d, and swabs taken less than 14days (21days for children) of vaccination will not be included in descriptive analyses.

***Flu TNCC***

For analysis of this test-negative case-control study, logistic regression will be used to calculate the unadjusted odds ratios for influenza vaccination in cases compared to controls, with a 95% confidence interval. This will be used to calculate an unadjusted VE as unadjusted VE=1-OR.

Logistic regression will be used to calculate the odds ratio for vaccination, adjusted for week of swab, scheme and age group, and additionally for relevant characteristics which change the vaccine effect by ≥1% (including risk status and sex). This will be used to calculate the adjusted VE. This will be undertaken for all cases/controls in the elderly, working-age adults and children separately, and in relevant subgroup analyses.

***Missing data***

We will aim to use multiple imputation methods for our main analysis where possible, rather than exclude data. However, data on vaccination status, age group and date of swab must be complete for inclusion in the study.

*Multiple imputation for missing risk status*

Multiple imputation methods will be used for missing data on risk status, taking into account patient age, and vaccination history and age eligibility.

*Multiple imputation for missing onset date*

Multiple imputation will be used where onset dates are missing. An interaction term between vaccination status and indicator for swab within 0-7days of onset will fitted (the sample size needs to remain consistent). Where onset is missing this indicator will be imputed based on the proportion of samples with known onset falling within the 7day window, taking into account positivity for flu or other viruses and patient age.

*Multiple imputation for missing vaccination date*

Where an adult patient has indicated that they were vaccinated at the time of the swab, but no vaccination date was given, a hot-deck imputation approach will be used. Vaccination dates will be sampled from other vaccinated patients within the same broad age cohort (18-64 or 65+) and swabbed within the same week. The imputed vaccination dates will be used to assign patients as within 1-13 days of vaccination or fully vaccinated.

In secondary analyses by vaccine type, a category for unknown vaccine type will be included. (Since vaccine type is almost universally missing for patients whose vaccination date is missing, the above imputation approach for vaccination dates need not be taken for analyses of vaccine type).

***Sensitivity analyses***

1. Exclusion of individuals with unknown onset date and missing data items.
2. Restriction to patients with Asthma/COPD exacerbation in RCGP data only, and/or exploration of indicators for swabbing (ARI/ILI/COVID-like illness).
3. Inclusion of SARS-CoV-2 positive cases, with/without adjustment for Autumn 2023 COVID-19 vaccination status.

***Sample size***

All eligible patients will be included. Expected cell counts of vaccinated cases, vaccinated controls, unvaccinated cases, unvaccinated controls will be calculated. If any expected cell count is below 10, the sample size is deemed too small to produce reliable estimates and VE will not be published.

#### 4) Dissemination of results

First V/E estimates (intra-seasonal) using the test-negative design will be disseminated among the research team and to WHO as part of the GIVE report early during the influenza season (early Jan of the influenza season) followed by the end of season analysis. We would envisage papers to be written at the end of the season following presentation of results at JCVI and VEBIS.

#### *5) Ethical approval*

The collection of the clinical data accords with routine practice. The analysis of swab forms according to positivity is currently undertaken as part of the routine assessment of the virological swabbing programme. The swabs are taken to assist clinical management.

UK public health agencies have permission to process patient confidential information for national surveillance of communicable diseases under: Regulation 3 of the Health Service Regulation 2002 for England, the Public Health (Scotland) Act 2008 and the NHS Scotland Act 1978 for Scotland, and the Public Health Wales National Health Service Trust (Establishment) Order 2009 for Wales. The work was reviewed by the UKHSA Research Ethics and Governance Group who confirmed the work was covered by Regulation 3 of The Health Service (Control of Patient Information) Regulations 2002, therefore specific ethical approval was not necessary. This review was also undertaken in each UK country.

### 6) Logistical aspects

The work described here is part of an ongoing programme which will continue according to funding availability.

#### 6.1 Study teams

**UKHSA**

Conall Watson, Heather Whitaker, Nick Andrews, Praveen Sebastian-Pillai, Katja Hoschler, Maria Zambon, Katie Hassell, Beatrix Kele

**Public Health Scotland**

Rory Gunson, Chris Robertson, Kimberly Marsh, Josie Evans, Mark Hamilton

**Public Health Wales**

Simon Cottrell, Catherine Moore, Christopher Williams, Jana Zitha, Panoraia Kalapotharakou, Sean Morgans, Anastazia Couzens, Kathleen Pheasant, Tim Jones

**University of Oxford / RCGP RSC**

Simon de Lusignan, Sneha Anand, Rosalind Goudie, Cecilia Okusi, Elizabeth Button, Vanashree Sexton, Jack Macartney, Timea Suli, Rashmi Wimalaratna, Rachel Byford, Gavin Jamie, Victoria Tzortziou-Brown, Utkarsh Agrawal
